# Spondyloarthritis, acute anterior uveitis, and Crohn’s disease have both shared and distinct gut microbiota

**DOI:** 10.1101/2022.05.13.22275044

**Authors:** Morgan Essex, Valeria Rios Rodriguez, Judith Rademacher, Fabian Proft, Ulrike Löber, Lajos Markó, Uwe Pleyer, Till Strowig, Jérémy Marchand, Jennifer A. Kirwan, Britta Siegmund, Sofia Kirke Forslund, Denis Poddubnyy

**Affiliations:** Experimental and Clinical Research Center (ECRC), a cooperation of the Max-Delbrück Center and Charité–Universitätsmedizin, Berlin, Germany; Max-Delbrück Center for Molecular Medicine in the Helmholtz Association (MDC), Berlin, Germany; Charité–Universitätsmedizin Berlin, a corporate member of Freie Universität Berlin and Humboldt–Universität zu Berlin, Berlin, Germany; Department of Gastroenterology, Infectious Diseases and Rheumatology, Campus Benjamin Franklin, Charité–Universitätsmedizin Berlin; Department of Ophthalmology, Campus Virchow, Charité–Universitätsmedizin Berlin; Metabolomics Platform, Berlin Institute of Health at Charité – Universitätsmedizin Berlin, Berlin, Germany; German Center for Cardiovascular Research (DZHK), partner site Berlin; Department of Microbial Immune Regulation in the Helmholtz Center for Infection Research, Braunschweig, Germany; Cluster of Excellence RESIST (EXC 2155), Hannover Medical School, Hannover, Germany; Centre for Individualised Infection Medicine (CiiM), a joint venture between the Helmholtz-Centre for Infection Research (HZI) and the Hannover Medical School (MHH), Hannover, Germany; University of Nottingham School of Veterinary Medicine and Science, Loughborough, UK; Structural and Computational Biology Unit, EMBL, Heidelberg, Germany; Department of Epidemiology, German Rheumatism Research Center (DRFZ), Berlin, Germany

**Keywords:** Gut microbiome, HLA-B27, Spondyloarthritis, Crohn’s Disease, Uveitis, Immune-mediated inflammatory disease

## Abstract

**Objectives:** Spondyloarthritis (SpA) is a group of immune-mediated diseases highly concomitant with non-musculoskeletal inflammatory disorders, such as acute anterior uveitis (AAU) and Crohn’s disease (CD). The gut microbiome represents a promising avenue to elucidate shared and distinct underlying pathophysiology.

**Method:** We performed 16S rRNA sequencing on stool samples of 277 patients (72 CD, 103 AAU, and 102 SpA) included in the German Spondyloarthritis Inception Cohort (GESPIC) and 62 back pain controls without any inflammatory disorder. Discriminatory statistical methods were used to disentangle microbial disease signals from one another and a wide range of potential confounders. Patients were naïve to or had not received treatment with biological disease-modifying anti-rheumatic drugs for at least three months before enrollment, providing a better approximation of a true baseline disease signal.

**Results:** We identified a shared, immune-mediated disease signal represented by low abundances of Lachnospiraceae taxa relative to controls, most notably *Fusicatenibacter*, which partially mediated higher serum CRP levels and was most abundant in controls receiving NSAID monotherapy. Patients with SpA drove an enrichment of *Collinsella*, while HLA-B27+ individuals displayed enriched *Faecalibacterium*. CD patients had higher abundances of a *Ruminococcus* taxon, and previous csDMARD therapy was associated with increased *Akkermansia*.

**Conclusion:** Our work supports the existence of a common gut dysbiosis in SpA and related inflammatory pathologies. We reveal shared and disease-specific microbial associations and potential mediators of disease activity. Validation studies are needed to clarify the role of *Fusicatenibacter* in gut-joint inflammation, and metagenomic resolution is needed to understand the relationship between *Faecalibacterium* commensals and HLA-B27.

## Introduction

The human gut microbiome is a largely symbiotic, complex ecosystem of microorganisms residing on the intestinal mucosal surface. Bacterial microbiome members, mainly of the Firmicutes and Bacteroidetes phyla, have been more widely studied than viral or fungal members, and confer many beneficial metabolic and immunological functions upon the host^1^. A dysbiotic microbiota composition is broadly defined as an imbalance between symbionts and pathobionts which reduces the resistance and resilience of the microbial gut ecosystem^2^. In a persistent dysbiotic state, physiological conditions such as epithelial barrier integrity may become compromised and increase intestinal permeability. This “leaky gut” phenomenon is thought to drive the inflammation characteristic of several immune-mediated diseases^3^.

Spondyloarthritis (SpA) refers to one such group of immune-mediated inflammatory diseases with a complex clinical spectrum in genetically-susceptible individuals. An estimated 50-75% of all SpA patients and as many as 90% of radiographic axial SpA patients carry the human leukocyte antigen (HLA)-B27 gene, making the association one of the strongest ever reported between an HLA allele and a disease^4^. The clinical SpA features include inflammation of the axial skeleton and extra-musculoskeletal manifestations such as psoriasis, acute anterior uveitis (AAU), and inflammatory bowel diseases (IBD), both Crohn’s disease (CD) and ulcerative colitis (UC)^5^. Up to 45% of SpA patients present with one or more extra-musculoskeletal manifestations in the course of their disease (around 33% with AAU and up to 15% with IBD), and around 20% of IBD patients and 40% of AAU patients eventually develop SpA^6–8^.

These diseases have a well-documented epidemiological association, but the underlying pathophysiological mechanisms are not yet fully understood despite decades of research. It was postulated more than 30 years ago that reactive arthritis (in the SpA disease family and sharing the genetic link to HLA-B27) could be triggered by antecedent gastrointestinal infection, after bacterial lipopolysaccharides were isolated from synovial fluid^9^. Also more than 25 years ago it was demonstrated that HLA-B27 transgenic rats, which spontaneously develop IBD and SpA pathologies, did not develop disease in a germ-free environment^10^. Subsequent gastrointestinal colonization with a few commensals was sufficient to trigger arthritis and colitis^11^, suggesting a causal role of the microbiome in (shared) pathogenesis. Despite the high co-occurrence of these diseases, most human microbiome studies have focused on bacterial alterations in SpA, CD, and AAU individually without exploring their concomitance. Cross-disease comparisons and meta-analyses have revealed that nearly half of microbial associations observed across diverse pathologies may not be specific, but rather part of a shared, more general disease signal^12,13^. Furthermore, it is increasingly clear that medication regimens exert a profound impact on microbiome composition^14–18^, and that studies which fail to account for treatment and disease concomitance are very likely to suffer from confounding and spurious associations with disease states. In our study, we aimed to characterize a robust shared microbiota among SpA, AAU, and CD for the first time in a large human cohort, and to further resolve relevant phenotypic and covariate associations therein.

## Methods

### Patient and public involvement

Neither patients nor the public were involved in the design, conduct, reporting, or dissemination plans of our research.

### Patient inclusion criteria

The German Spondyloarthritis Inception Cohort (GESPIC) is an ongoing prospective cohort initiated to study the course and long-term outcomes of SpA across its whole spectrum of clinical presentation, including but not limited to gut microbiome composition. In this study we cross-sectionally analyzed only the baseline data. Serum, stool, and peripheral blood mononuclear cell (PBMC) samples, demographic information, and clinical characteristics were collected.

Since September 2015, patients have been recruited in three different arms depending on their main condition: **1)** established radiographic axSpA: patients were required to fulfill the modified New York criteria and be eligible to start biological disease-modifying anti-rheumatic drug (bDMARD) therapy, by presenting high disease activity (BASDAI ≥ 4 and/or ASDAS ≥ 2.1) despite previous treatment with nonsteroidal anti-inflammatory drugs; **2)** Crohn’s disease^19^: patients were classified according to the Montreal classification including location and behavior of CD, and had been recently diagnosed; **3)** acute anterior uveitis^20^: patients with non-infectious AAU diagnosed by an ophthalmologist. CD and AAU patients were enrolled regardless of musculoskeletal symptoms and an experienced rheumatologist was responsible for the final diagnosis of SpA / no SpA for patients included in these cohorts. All cohorts were approved by the ethical committee (Charité-Universitätsmedizin Berlin, Berlin, Germany). All patients enrolled were at least 18 years of age and gave their written informed consent. Patients included in this analysis were naïve to or had not received treatment with bDMARDs for at least three months before the enrollment in the study, nor had they received systemic antibiotics for at least one month prior to their baseline stool sample. There were no other restrictions concerning therapy. Control individuals were selected from the Optiref study^21^, which consisted of patients with chronic back pain who went through a standardized rheumatologic examination where the diagnosis of SpA was ruled out. Individuals with CD, AAU or psoriasis were excluded from the control group.

### 16S rRNA amplicon sequencing

After storage at -80°C, fecal samples were defrosted on ice. Aliquots of fecal material (1ml) resuspended in RNALater were centrifuged and washed once with water to remove excess fixative and salt. Then, DNA was isolated using the ZymoBIOMICS DNA Miniprep Kit (Zymo Research) according to the manufacturer’s instructions. Bead Beating was performed four times for 5 minutes each. Amplification of the V4 region (F515/R806) of the 16S rRNA gene was performed according to previously described protocols^22^ across two separate experiments. Briefly, 25 ng of DNA were used per PCR reaction (30 μl). The PCR amplification was performed using Q5 polymerase (NEB Biolabs). The PCR conditions consisted of initial denaturation for 30s at 98°C, followed by 25 cycles (10s at 98°C, 20s at 55°C, and 20s at 72°C). Each sample was amplified in triplicates and subsequently pooled. After normalization PCR amplicons were sequenced on an Illumina MiSeq platform (PE300).

### Taxonomic profiling and preprocessing

Computational analysis was carried out in R (v4.0.3). Demultiplexed reads were analyzed using the DADA2 pipeline (v1.25.2) and taxonomic assignment was based on the SILVA rRNA database (v138.1) at 80% bootstrapped confidence. A total of 2118 unique bacterial ASVs were identified across 435 samples, annotated at the genus- or family-level. The *DECIPHER* package was used for multiple sequence alignment and the *phangorn* package was used to build a phylogenetic tree. Samples failing to meet inclusion criteria, lacking important clinical metadata, or not passing quality control (i.e. library sizes <5000 and >38K total raw reads) were excluded from further analysis (**SFig. 1A**), resulting in 339 samples. Libraries were rarefied to 5000 reads to account for variability in sequencing depth, resulting in 2066 ASVs present in at least one sample. The *phyloseq::tax_glom()* function was implemented in a custom manner that binned ASVs at the lowest taxonomic annotation available, either genus (70% of ASVs) or family (remaining 30%), and separately according to phyla (100% of ASVs). After total-sum scaling (TSS) normalization, three datasets: (i) ASV relative abundances, (ii) genus- or family-level relative abundances, and (iii) phylum-level relative abundances, which could be further processed (i.e. filtered) as needed.

### Alpha and beta diversity analysis and enterotyping

The *phyloseq::estimate_richness()* function was applied to raw sample counts in order to calculate the Shannon entropy as a measure of alpha diversity (within-sample variation). To assess beta diversity (between-sample taxonomic variation), the *vegan::vegdist()* function was applied to rarefied ASV profiles (after TSS normalization) to calculate pairwise Bray-Curtis dissimilarity matrices and the *stats::cmdscale()* function was used to perform a principal coordinates ordination analysis (PCoA). The *DirichletMultinomial* R package was used to enterotype samples from rarefied counts binned at the genus- or family-level, according to the procedure from Holmes, Harris, and Quince^23^.

### Differential abundance analysis

ASVs not present in more than 5% of samples or displaying mean relative abundances less than 1e-05 across all samples were filtered out, resulting in n=442 ASVs and n=123 binned higher-level taxa (annotated at either genus- or family-level, see **Supplementary Table 1**) for univariate statistical testing and confounder analysis. To examine a potential shared disease signature, all n=277 disease samples (“GESPIC”) were pooled and compared to the controls in an integrated differential abundance and association testing procedure. Samples were also grouped according to phenotype and phenotypes with n>40 samples per group (i.e. SpA-only, CD-only, AAU-only, SpA+AAU) were subjected to this procedure to resolve phenotype-specific associations. The *phyloseq* and *ggtree* packages were used to visualize the phylogenetic information of ASVs.

Custom linear models were built for all n=565 taxa using the *lm* function in base R with the formula *relative_abundance* ∼ *disease_status * sequencing_experiment* to account for (un)known technical variation. Model parameters were extracted, tested for significance using the default Wald method, adjusted for multiple testing (Benjamini-Hochberg procedure), and bootstrapped to obtain 95% confidence intervals using the *parameters::model_parameters()* function. As a robustness check, all taxa were tested for significant differential abundance using a blocked Wilcoxon permutation test (*coin::wilcox_test()* function with the formula *relative_abundance* ∼ *disease_status* | *sequencing_experiment*), and raw *P* values were adjusted for multiple testing.

### Confounder and mediation analysis

Keeping the same sets of taxa and phenotypic groups described above, non-parametric effect sizes (Cliff’s delta and the Spearman correlation) were calculated for each possible pairing of taxonomic and clinical features (see **Supplementary Table 2** for a complete list), then tested for significance independent of a disease signal using blocked Wilcoxon or Spearman tests from the *coin* package (formula *relative_abundance* ∼ *covariate* | *disease_status*), and adjusted for multiple testing (Benjamini-Hochberg procedure).

The *mdt_simple()* function from the *JSmediation* package was used as described in Yzerbyt *et al*.^*24*^ to test specific causal mediation hypotheses. This approach used linear regression to estimate the paths (a, b, c, and c’) along a possible causal pathway involving three variables. The joint significance of these paths was assessed to determine if mediation was present, and if so what the direct and indirect contributions within the pathway were. To lower the burden of multiple testing, only configurations with significantly disease-associated taxa (as in **Fig. 2**) and covariates emerging from the above testing were considered.

Further linear models were built with variables tracking current and prior medication intake (see **Supplementary Table 3**, detailed therapies in **Table 1** were not used) to estimate the impact of specific drug therapies on taxon abundances. Instead of using the case-control groupings above, samples were first grouped according to disease phenotype (groups with <40 samples were not analyzed), such that disease status was not a factor, and the formula *relative_abundance* ∼ *intake_status * sequencing_experiment* was used to test specific disease-drug differential abundance hypotheses (e.g. conventional synthetic (cs)DMARD use, which was only relevant in CD patients). In the group of all patients examining previous antibiotic use, the formula *relative_abundance ∼ disease_status * sequencing_experiment + intake_status* was used; in the control subset examining NSAID use, simple models (*relative_abundance ∼ intake_status*) were used as no other factors were relevant.

**Table 1:** Clinical baseline characteristics for each cohort (CD, axSpA and AAU) and controls. Calculated *P* values represent chi-square or Kruskal-Wallis (denoted with §) tests between all n=4 groups or disease cohorts only (denoted with #). AAU, anterior acute uveitis; (ax)SpA, (axial) spondyloarthritis; CD, Crohn’s disease; HLA-B27, human leukocyte antigen B27; BMI, body mass index; CRP, C-reactive protein; DMARD, disease-modifying antirheumatic drug; bDMARD, biological DMARD; csDMARD, conventional/synthetic DMARD; NSAID, non-steroidal anti-inflammatory drug; PPI, proton pump inhibitor; SD, standard deviation.

|  | CD<br>n = 72 | AAU<br>n = 103 | axSpA<br>n = 102 | Controls<br>n = 62 | P value |
| --- | --- | --- | --- | --- | --- |
| Age in years, mean ± SD | 37.4 ± 12.7 | 42.8 ± 13.1 | 36.9 ± 10.6 | 38.4 ± 10.4 | 0.003<sup>§</sup> |
| Male sex, n (%) | 34 (47.2) | 48 (46.6) | 65 (63.7) | 26 (41.9) | 0.015 |
| HLA-B27 positive, n (%) | 6 (8.3) | 81 (78.6) | 89 (87.3) | 5 (8.1) | <0.001 |
| BMI in kg/m<sup>2</sup>, mean ± SD | 24.4 ± 4.6 | 24.7 ± 5.7 | 25.1 ± 4.2 | 25.9 ± 5.0 | 0.153<sup>§</sup> |
| Smoking status: current smoker, n (%) | 24 (33.3) | 22 (21.4) | 38 (37.3) | 11 (17.7) | 0.02 |
| Alcohol intake, g/day, mean ± SD | 3.6 ± 6.3 | 2.1 ± 3.6 | 2.1 ± 4.2 | - | 0.14<sup>#</sup> |
| SpA, n (%) | 12 (16.7) | 55 (53.4) | 102 (100) | 0 | <0.001 |
| Uveitis ever, n (%) | 11 (15.3) | 103 (100) | 22 (21.6) | 0 | <0.001 |
| Psoriasis ever, n (%) | 4 (5.6) | 10 (9.7) | 17 (16.7) | 0 | 0.002 |
| Crohn's disease ever, n (%) | 72 (100) | 1 (1.0) | 7 (6.9) | 0 | <0.001 |
| CRP mg/l, mean ± SD | 11.0 ± 26.9 | 4.1 ± 7.3 | 14.0 ± 18.7 | 1.2 ± 1.8 | <0.001<sup>§</sup> |
| Current NSAID treatment, n (%) | 18 (25.0) | 36 (35.0) | 98 (96.1) | 39 (62.9) | <0.001 |
| Current PPI treatment, n (%) | 6 (8.3) | - | 40 (39.2) | - | - |
| Current systemic corticosteroid treatment, n (%) | 26 (36.1) | 17 (16.5) | 5 (4.9) | 0 | <0.001<sup>#</sup> |
| Current csDMARD treatment, n (%) | 33 (45.8) | 6 (5.8) | 3 (2.9) | - | <0.001<sup>#</sup> |
| Sulfasalazine, n (%) | 1 (1.4) | 1 (1.0) | 2 (2.0) | - | <0.001<sup>#</sup> |
| Methotrexate, n (%) | 0 | 5 (4.9) | 1 (1.0) | - | <0.001<sup>#</sup> |
| Mesalazine, n (%) | 10 (13.9) | 0 | 0 | - | <0.001<sup>#</sup> |
| Azathioprine, n (%) | 21 (29.2) | 0 | 0 | - | <0.001<sup>#</sup> |
| Naive to csDMARD treatment, n (%) | 23 (31.9) | 90 (88.2) | 98 (95.1) | - | <0.001<sup>#</sup> |
| Current bDMARD treatment, n (%) | 0 | 0 | 0 | - | - |
| Naive to bDMARD treatment, n (%) | 70 (97.2) | 95 (92.2) | 81 (79.4) | - | <0.001<sup>#</sup> |

## Results

### Clinical presentation of SpA, AAU, and CD cohorts

Demographic and clinical characteristics of our 277 patients (72 CD, 103 AAU, and 102 axSpA patients) and 62 back pain controls are detailed in Table 1. More than half of the AAU cohort and one fifth of the CD cohort presented with predominantly axial SpA, with just five cases from the CD and three from the AAU cohorts diagnosed as exclusively peripheral SpA. A large majority of AAU and axSpA cohort patients carried HLA-B27 while less than 10% of individuals in the CD and control groups did (**Table 1** and **SFig. 1B**). Over 90% of all patients were naïve to biologic disease-modifying anti-rheumatic drugs (bDMARDs, or biologics). In terms of disease activity, only patients in the SpA cohort showed clearly high systemic inflammatory activity, with mean CRP levels of 14.0mg/l and Ankylosing Spondylitis Disease Activity Score (ASDAS) of 3.5±0.8. Patients in the CD cohort had relatively inactive disease with a mean Harvey-Bradshaw Index (HBI) of 3.3±3.9, and patients in the AAU cohort had mean CRP levels of 4.1mg/l, although 45 (43.7%) had an active episode of anterior uveitis at the time of enrollment.

### High-level variation and taxonomic diversity of gut microbiota

We performed 16S rRNA sequencing and taxonomically profiled a total of 339 stool samples (277 patients and 62 disease-negative back pain controls). At the phylum level our disease cohorts were dominated by Firmicutes, followed by Bacteroidota, Actinobacteriota and Proteobacteria (**SFig. 2A, 2B**). At the genus level, we observed substantially more variation between individuals in all cohorts, with *Bacteroides, Prevotella*, and *Faecalibacterium* comprising the dominant genera in each cohort (**SFig. 2A, 2C**). Notably, back pain controls and patients from the axSpA and AAU cohorts had more *Prevotella-*dominant individuals on average than did the CD cohort, which had the highest proportion of *Bacteroides*-dominant individuals (**SFig. 2B, 2C**). Patients with CD phenotypes had the lowest alpha diversities (**Fig. 1A, 1C**), but beta diversity did not appear to correlate with disease phenotype (**Fig. 1B**) or technical variation due to the sequencing experiment (**Fig. 1D**).

**Figure 1:**
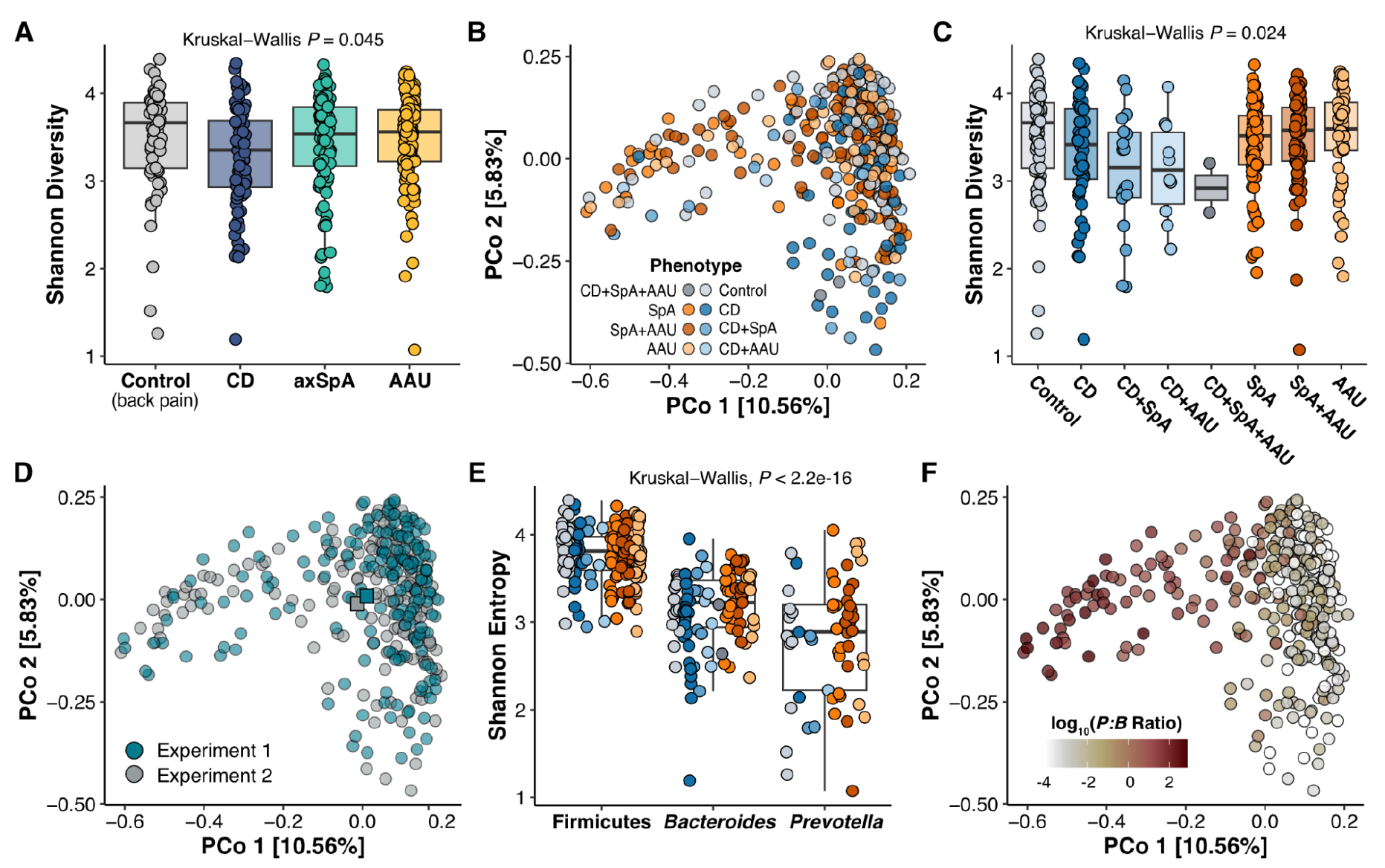
CD phenotypes display lowest alpha diversity and beta diversity is correlated with the ratio of *Prevotella:Bacteroides*. The Shannon entropy was selected to capture the alpha diversity and pairwise Bray-Curtis dissimilarities were used for a principal coordinates ordination analysis (PCoA) to evaluate beta diversity (see **Methods**). **A)** Alpha diversity grouped by primary cohort. **B)** Beta diversity, samples colored according to phenotype. **C)** Alpha diversity grouped by phenotype. **D)** Beta diversity, samples colored according to sequencing experiment with the spatial means of each group depicted (squares). **E)** Alpha diversity grouped by phenotype and enterotype (see **Methods** and **SFig. 2**). **F)** Beta diversity, samples colored according to the *Prevotella-*to-*Bacteroides* ratio^25^ which correlated with the first projected ordination axis (Pearson’s *r* = -0.8, *P*<0.001), accounting for up to 10% of between-sample variation and indicating an inverse relationship between these taxa. AAU, acute anterior uveitis; CD, Crohn’s disease; (ax)SpA, (axial) spondyloarthritis.

**Figure 2:**
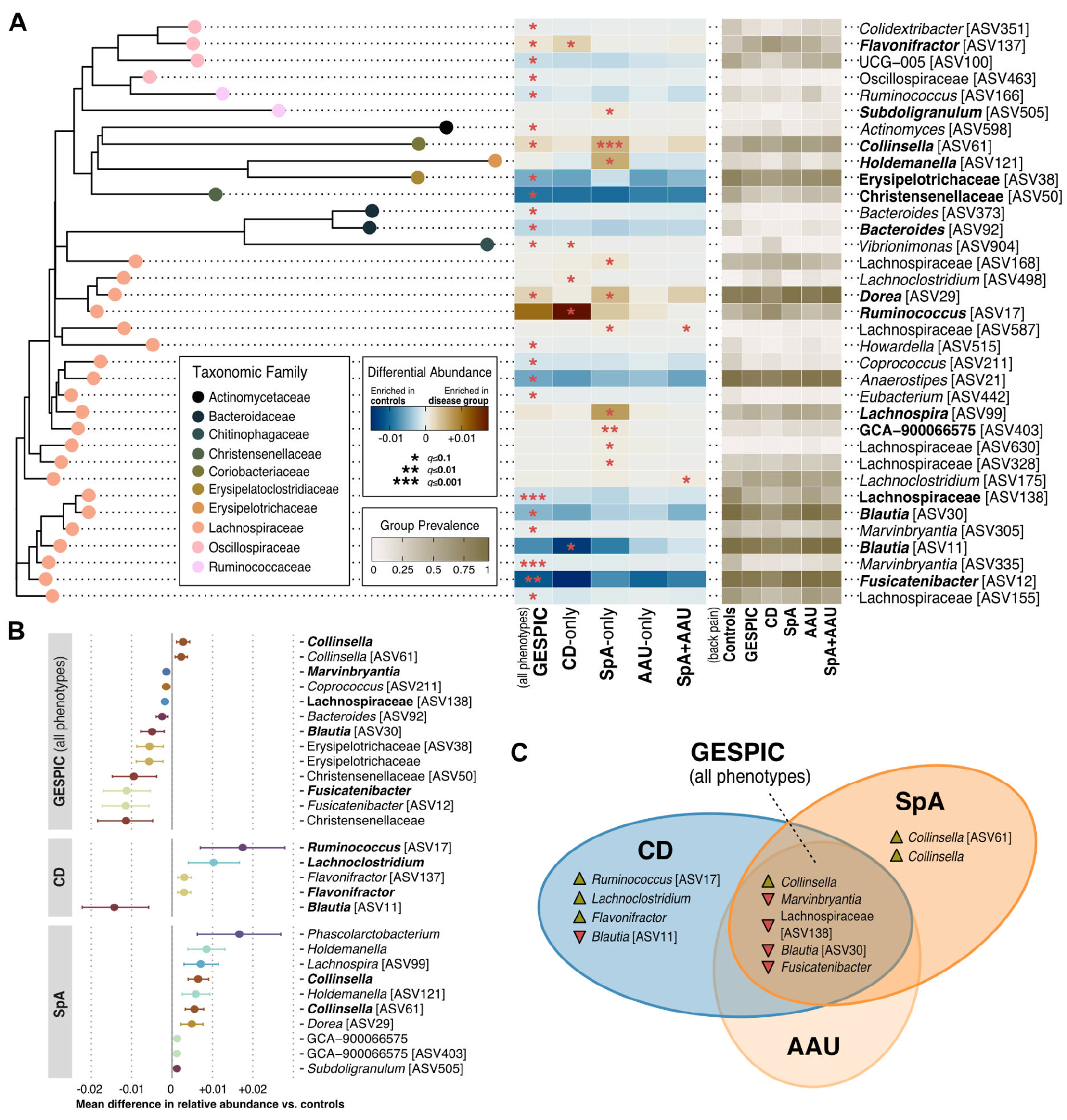
Immune-mediated disease phenotypes share a depletion of Lachnospiraceae taxa and an SpA-driven enrichment in *Collinsella*. Disease groups consisted of patients without concomitant disease unless explicitly stated (e.g. SpA+AAU); GESPIC refers to the group of all n=277 patients. Linear models were built for ASVs (n=442) and their genus- or family-level taxonomic bins (n=123), containing an interaction term to account for technical variation (see **Methods**). **A)** Phylogenetic relationships, regression coefficients, and prevalence of ASVs significantly associated with one or more disease states. Bolded taxa had an adjusted *q*<0.05 and an absolute coefficient estimate >0.001 (corresponding to a mean differential abundance between disease and control groups of .1%). The x-axis broadly represents evolutionary distance based on the 16S V4 amplicons. **B)** Regression coefficients for the bolded taxa in A, plus any bins which were significantly differentially abundant, colored by genus or family with 95% bootstrapped confidence intervals shown. Bolded taxa were significant using a blocked Wilcoxon test at a *q*<0.05 cutoff. **C)** Venn diagram containing the bolded taxa from B, summarizing the most robust associations. ASV, amplicon sequence variant; GESPIC, German Spondyloarthritis Inception Cohort; CD, Crohn’s disease; SpA, spondyloarthritis; AAU, acute anterior uveitis.

### Taxonomic associations with immune-mediated inflammatory disease states

To uncover taxa potentially mediating the concomitance of SpA, CD, and AAU, we performed a differential abundance analysis with disease-negative controls. We observed significantly lower mean relative abundances of several Firmicutes in the disease group as a whole, mainly from ASVs belonging to the Lachnospiraceae family (**Fig. 2A**). Of these, *Fusicatenibacter* [ASV12], *Anaerostipes* [ASV21], and *Blautia* [ASV30] were among the most prevalent. One ASV from the Christensenellaceae family (a health-associated^26^ taxon with known heritability^27^) displayed the strongest overall depletion in the GESPIC group relative to the control group; when all seven ASVs in our data belonging to this family were binned, the magnitude of the depletion increased (**Fig. 2B**). Similar behavior was observed for *Fusicatenibacter, Marvinbryantia*, and Erysipelotrichaceae ASVs, but not for *Bacteroides, Blautia*, or Lachnospiraceae ASVs, which were, in contrast, not significantly differentially abundant at higher taxonomic levels. GESPIC patients also had significantly higher abundances of *Collinsella* and *Flavonifractor*, both of which tracked closely with single ASVs and were ultimately found to be driven by the SpA and CD phenotypes, respectively (**Fig. 2C**). In addition to *Collinsella*, the SpA group was enriched in several Lachnospiraceae ASVs, *Subdoligranulum* [ASV505], and *Holdemanella* [ASV121] (**Fig. 2A**), few of which were significant when binned (**Fig. 2B**). Similarly, the CD group was strongly enriched in a *Ruminococcus* ASV and strongly depleted in a *Blautia* ASV (**Fig. 2A**), neither of which were significant at their respective genus levels (**Fig. 2B**). In contrast, *Phascolarctobacterium* and *Lachnoclostridium* reached significance when binned, but not at the ASV level (in the SpA and CD groups, respectively). There were no ASVs or higher-level taxa significantly associated with the AAU phenotype and only two weakly enriched Lachnospiraceae ASVs in the SpA+AAU phenotype; however, adjusted mean differential abundances for these groups mostly followed the pattern of the other case-control comparisons (**Fig. 2A**). Taken together, our findings broadly reflect the known taxonomic diversity of the Lachnospiraceae family^28,29^ at the clinical level, and reveal shared and disease-specific taxonomic signatures ranging from ASV to family resolution.

### Impact of drug therapies in GESPIC patient microbiota

To clarify the disease-associated microbiota signals, we examined a wide range of clinically-relevant factors. We found no significant associations between any taxa and age, sex, or BMI, indicating our disease signals were unlikely to be confounded or mediated by demographic factors. Medication intake has been found to explain more variation in microbiome composition than disease status alone^16,17^ and is therefore an important factor to consider in association studies. In contrast to much of the literature^30–33^, no individuals in our study were taking bDMARDs at the time of sampling and a vast majority were naive to any biologic therapies, allowing us a better approximation of a true baseline disease signal. However, 70% (including back pain controls) were taking at least one glucocorticoid, csDMARD, or NSAID, and 22% were taking two concurrently (**Supplementary Table 3**).

As >95% of our SpA patients were taking NSAIDs, we could not examine the disease signal independent of this effect; however, 60% of our control group was taking NSAID monotherapy, which was associated with 1-2% more *Fusicatenibacter* [ASV12] and *Subdoligranulum* [ASV16] on average (**Fig. 3A**). Interestingly, these two genera specifically were found to predict good response to csDMARDs in rheumatoid arthritis patients^34^. 38% of our SpA-only patients were additionally receiving proton pump inhibitors, which was associated with an increase in *Phascolarctobacterium* (**Fig. 3B**), likely confounding that signal (**Fig. 2B**). No other disease-associated taxa appeared to be sensitive to the drugs we were able to test.

**Figure 3:**
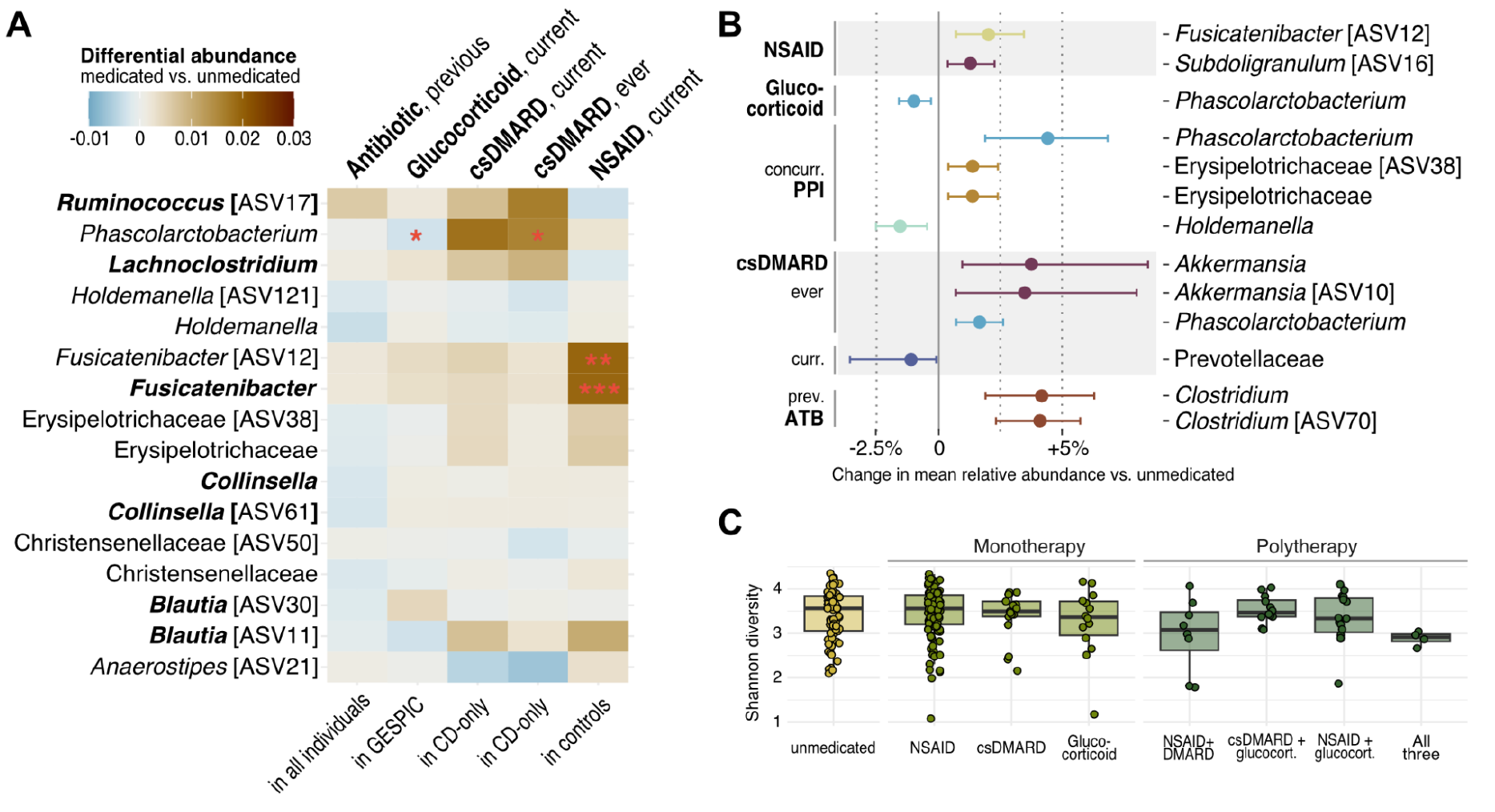
*Fusicatenibacter* is enriched in NSAID monotherapy, and *Akkermansia* is enriched in CD patients previously treated with csDMARDs. **A)** Linear regression coefficients for disease-associated taxa (**Fig. 2B**), estimating the differential abundance between medicated and unmedicated patients, either while adjusting for disease status and/or technical variation (two leftmost columns, see **Methods**), or resulting from a comparison within a specific subset of patients (specified underneath). Bolded taxa were most robustly associated with CD, SpA, or the shared signal (**Fig. 2C**). We had information tracking previous antibiotic use (>1 month and <3 months before sampling) for all individuals in our study. **B)** Regression coefficients from the same analyses in **A**, for any taxa with an absolute coefficient estimate >0.01 (corresponding to a mean difference between medicated and unmedicated groups of 1%), colored by genus or family with 95% bootstrapped confidence intervals shown. **C)** Alpha diversity among all n=262 patients for which information on current intake for all three drug therapies was available. ATB, antibiotic.

Nearly half our CD-only patients were on a form of csDMARD therapy at the time of sampling, which was associated with significantly less Prevotellaceae (**Fig. 3B**). When comparing csDMARD-naive CD patients to those that were currently or had previously received treatment, we found that treated individuals had increased *Phascolarctobacterium* and *Akkermansia* (**Fig. 3B**). Although species-level identification from amplicon data is a contentious practice we avoided here, it is worth noting that these individuals also had about 1% more of a *Ruminococcus* taxon [ASV17] identified as *R. gnavus*^*28,35*^, enriched in our CD patients and elsewhere (using metagenomic data) in treatment-resistant UC patients^36^, as well as SpA patients with concomitant CD^33^.

### Mediation of inflammation and disease activity by HLA-B27 and microbiota

Other clinically-relevant covariates included the presence or absence of the HLA-B27 antigen on immune cells, systemic inflammation (as measured by C-reactive protein, CRP), and disease activity scores (**Fig. 4A**). *Fusicatenibacter* was the taxa most strongly (negatively) correlated with CRP (**Fig. 4B**), and we estimated about 19% of the increase in CRP observed in our GESPIC group to be mediated by [the relative lack of] this taxon (**Fig. 4C**). Several *Subdoligranulum, Roseburia, Lachnospira*, and *Faecalibacterium* ASVs as well as the total (binned) *Faecalibacterium* abundance were significantly higher in HLA-B27+ individuals, whereas *Escherichia-Shigella* and *Blautia* [ASV83] abundances were significantly lower in those individuals (**Fig. 4D**).

**Figure 4:**
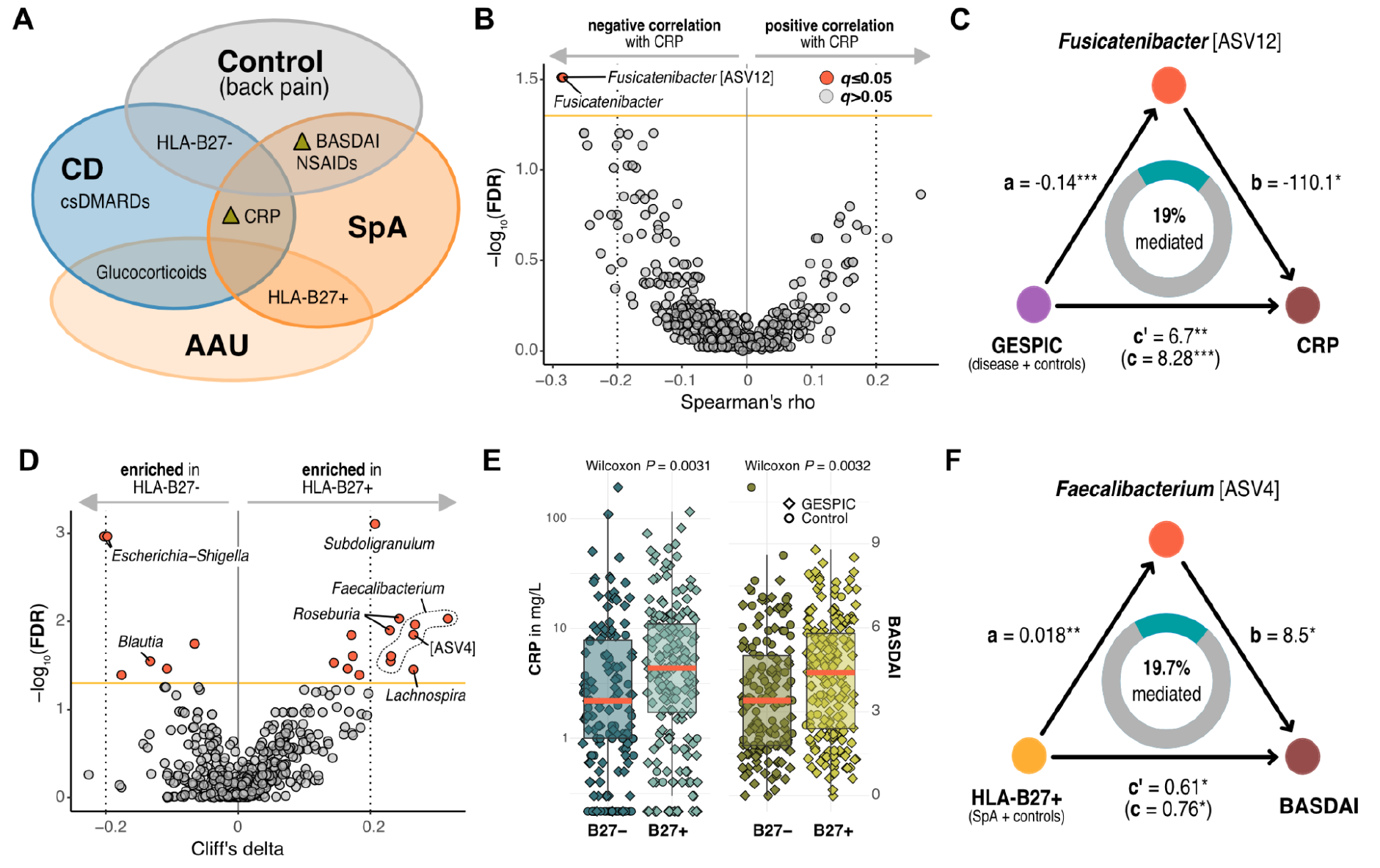
Decreased *Fusicatenibacter* partially mediates increased CRP in patients with SpA or CD, and increased *Faecalibacterium* partially mediates higher disease activity in HLA-B27+ SpA patients. **A)** Summary of **Table 1** highlighting clinical factors differentiating our phenotypes. **B)** Non-parametric effect sizes and significance tests showing serum CRP correlations independent of disease status for all n=585 taxa across all n=339 individuals in our study. **C)** Mediation analysis where *c* represents the total effect of disease presence on serum CRP, and the sum of both direct (*c’*) and indirect (*a*x*b*) paths, with the effect and significance of each component (*a, b, c, c’*) illustrated (see **Methods**). When accounting for (decreased) *Fusicatenibacter* abundances, serum CRP estimates were (significantly) further increased, from 6.7 mg/L to 8.28 mg/L in the GESPIC group compared to controls. \**q*<0.1, \*\**q*<0.01, \*\*\**q*<0.001. **D)** Similar to **B**, with HLA-B27. **E)** HLA-B27+ individuals (mostly SpA and AAU) had increased inflammation and disease activity. **F)** Mediation analysis where *c* represents the total effect of HLA-B27 expression status on BASDAI scores and *c’* the indirect effect accounting for the enriched *Faecalibacterium* [ASV4] observed in HLA-B27+. BASDAI, Bath Ankylosing Spondylitis Activity Index.

Since our disease phenotypes were significantly different in their CRP levels and expression of HLA-B27 (**Fig. 4A**), we hypothesized that some of our shared disease-associated taxa or HLA- and inflammation-associated taxa might serve as mediators. Regardless of group, none of the taxa we considered (nor summary features such as taxonomic richness and alpha diversity) were found to mediate the increase in CRP observed in HLA-B27+ individuals in our study (**Fig. 4E**). However, in the group of SpA-only patients and back pain controls (who had higher BASDAI scores on average than patients from the CD or AAU cohorts but few HLA-B27+ individuals), we found that *Faecalibacterium* [ASV4] mediated about 20% of the (modestly) significantly increased disease activity in HLA-B27+ individuals (**Fig. 4F**).

### Pilot study of serum metabolites in immune-mediated pathologies

Lachnospiraceae taxa are among the main short-chain fatty acid (SCFA) producers in the gut^28,29^, and dysbiosis also appears to be tightly correlated with the local balance between Tregs and pro-inflammatory Th17 cells^37^, which respond to microbially-produced tryptophan metabolites^38^. We performed a small, targeted pilot study to quantify these metabolites in the serum (see **Supplementary Methods**), and observed higher circulating levels of SCFA in GESPIC patients relative to controls, although not statistically significant (**Fig. 5A**). This was surprising but consistent with work that correlated increased circulating SCFAs with gut permeability and dysbiosis in diabetics^39^. No metabolites correlated with disease-associated taxa, likely due to low statistical power. The kynurenine-to-tryptophan ratio (KYN/TRP, proposed as a metric of immune activation^40^) was positively correlated with CRP in disease groups (**Fig. 5B**), in line with a previous study which found anti-TNFα therapy in SpA to be effective at reducing both clinical markers^41^. Serum serotonin concentration, found elsewhere^42^ to stratify CD patients into disease activity categories better than both CRP and KYN/TRP, was also strongly negatively correlated with BASDAI scores in individuals with SpA (**Fig. 5C**).

**Figure 5:**
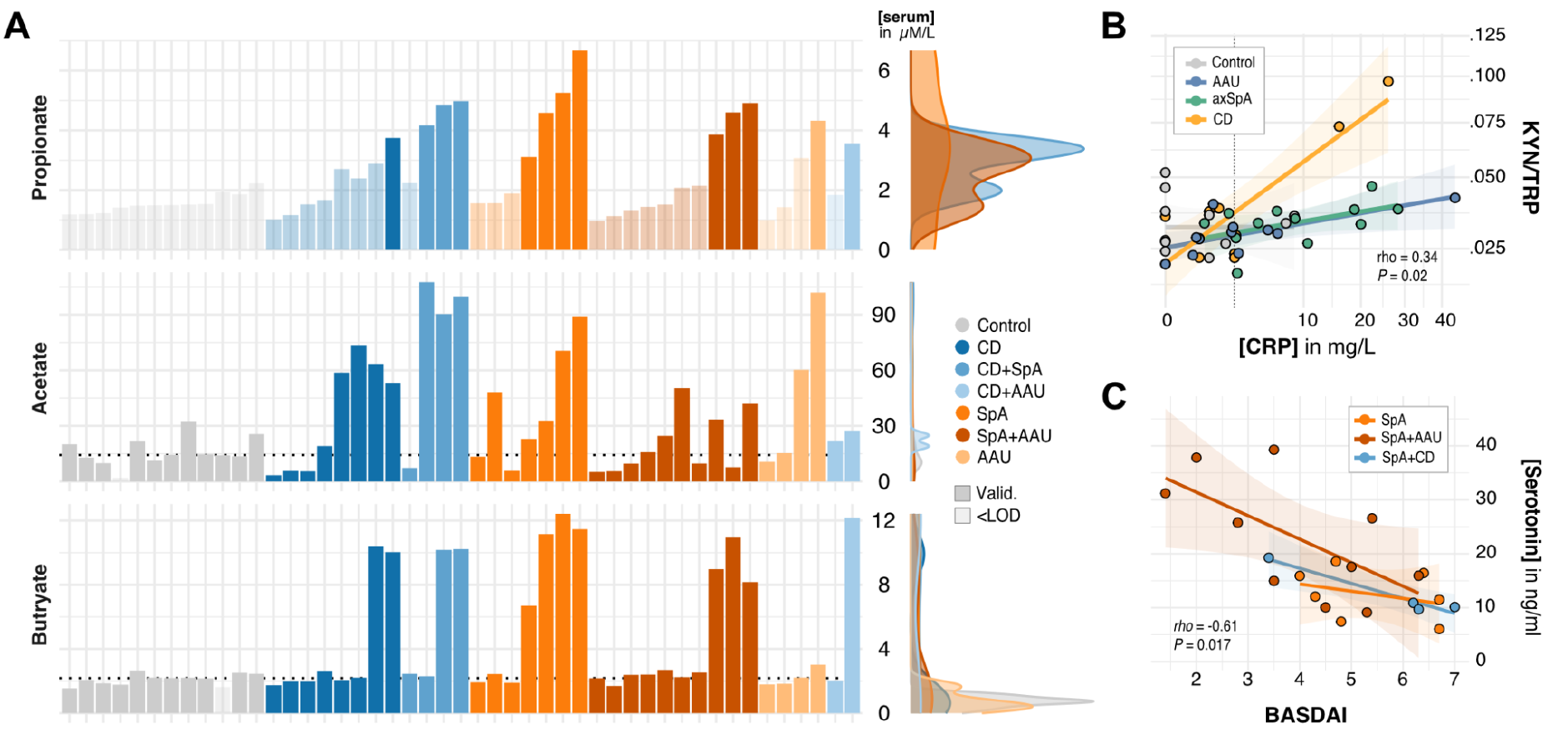
Serum serotonin and the kynurenine-to-tryptophan ratio correlate with disease activity scores and systemic inflammation. **A)** Concentrations of circulating short-chain fatty acids (SCFAs, in *μ*M/L), which were not significantly associated with any disease but appeared higher in disease patients (if available, median of the control group is shown as a dotted line). Some metabolites were below the limit of detection (<LOD), especially propionate, and thus not used for statistical analysis (see **Supplementary Methods**). Marginal density histograms include only valid measurements. **B)** Spearman correlations between systemic inflammation (CRP, in mg/L) and the kynurenine-to-tryptophan ratio (KYN/TRP), which were positive in disease cohorts, but not the control group (*rho* = -0.06). CRP values >5 mg/L (dotted line) implicate active disease. Both axes are square-root transformed to aid visualization. **C)** Spearman correlations between BASDAI scores and serum serotonin concentrations (in ng/ml). Overall Spearman correlation coefficients and *P*-values shown in B and C, blocked for groups shown in the respective legends. KYN, kynurenine; TRP, tryptophan.

## Discussion

CD, AAU and SpA share an established epidemiology, yet the pathophysiology underlying their concomitance remains unclear. We analyzed a large human cohort comprising all three diseases, and deeply explored the taxonomic composition of the gut microbiota with clinical covariates and disease concomitance for the first time. Our results showed a shared depletion of predominately Lachnospiraceae taxa, most notably *Fusicatenibacter*, which partially mediated increased CRP, and was most abundant in controls receiving NSAID monotherapy. SpA individuals had a robust enrichment of *Collinsella* relative to chronic back pain controls, and HLA-B27+ individuals (regardless of disease phenotype) displayed enriched *Faecalibacterium*.

It has been hypothesized elsewhere that HLA-B27 plays a causal role in SpA pathology^43^, and that it shapes the gut microbiota composition^4^. Our causal mediation analysis revealed that the increased *Faecalibacterium* [ASV4] we observed in HLA-B27+ patients partially mediated the increased BASDAI in those same patients. This intriguing result implies that this bacterium *contributes* to discomfort, pain, and fatigue perceived in SpA patients. This seems to contradict its known anti-inflammatory role in inflammatory bowel diseases^44^, yet aligns with similar HLA-B27 +/- microbiota comparisons in the SpA literature^45^ and our finding that abundances of this taxon were positively correlated with BASDAI scores (Spearman’s *rho* = 0.22, *q* = .009).

Although we did not observe a strong microbiota signal in our AAU-only patients, our results suggest that AAU patients with concomitant SpA (92% HLA-B27+) are more similar to SpA-only patients than disease-negative controls (**Fig. 2**). Association studies seeking to link the HLA-B27+ microbiota to an existing clinical understanding of these diseases^20,46^ should prioritize inclusion of non-SpA HLA-B27+ groups for more precise, powered comparisons – both from other diseases (i.e. AAU as we did here), and from disease-negative and genetically similar individuals, as in Berland *et al*.^*33*^. Our control group mirrored the European population and was mostly HLA-B27-, but had chronic back pain. Since half were receiving NSAID monotherapy, we were able to estimate the impact of this treatment on individual taxa and further strengthen our hypothesis that *Fusicatenibacter* is a key microbe mediating host gut-joint inflammation. This genus has only one known species, *F. saccharivorans* (identified as such in our data as ASV12), which alleviated colitis in a murine model and induced anti-inflammatory IL-10 production in lamina propria cells from UC patients^47^. *Blautia* emerged from our analyses as another genus likely to play a role in immune-mediated inflammatory pathologies^48,49^, although one that should perhaps be characterized with more genomic resolution than we had access to here^28^.

Restriction to amplicon sequencing data was a major limitation of our study, precluding further characterization important for understanding potential disease pathomechanisms. Yet, the ASVs analyzed here represent *de novo* sequences which more accurately capture taxonomic diversity and are more reproducible than OTU-based approaches^50^. Furthermore, the functional microbiota is intrinsically coupled to a set of discrete bacterial units (taxa) in an ecological context^51,52^, and while some functional differences indeed correlate with strain-level organisms or even nucleotides (as in probiotic therapies^53^ and bacterial SNPs^54^, respectively), higher-level bacterial taxonomies are still clinically useful to stratify disease patients and generate testable hypotheses in experimental or animal disease models.

For example, our SpA patients exhibited higher abundances of *Collinsella* (in line with previous results^55,56^), a genus which has elsewhere been shown to reduce the expression of enterocyte tight junction proteins *in vitro* (potentially contributing to gut leakage *in vivo*), as well as increase the production of pro-inflammatory IL-17A and transcription factor NFkB1^46^. This is relevant in SpA, where an excessive activation of IL-17A drives an expansion of Th17 cells, which further perpetuates IL-17A production^57^ (and thereby contributes to chronic inflammation). Similarly, colitis is associated with hyperproduction of Th17 cells, partly resulting from dysregulated NFκ-B activation responsible for inflammatory T cell differentiation^58^. Previous CD-focused work isolated a 15kDA microbial anti-inflammatory molecule from *F. prausnitzii* able to inhibit NFκ-B signaling *in vitro* and alleviate colitis in mice^59^; however, *Faecalibacterium* [ASV4] was *not* identified as this species in our data (although others in **Fig. 4D** were). Like *Blautia, Faecalibacterium* taxa appear to require at least metagenomic resolution to unravel their clinically-relevant properties^60^.

Here we presented the baseline cross-section of a prospective cohort examining SpA, CD, and AAU. Taken together, our results suggest there is much more to be uncovered about the immunomodulatory properties of certain bacteria in these epidemiologically-related pathologies, especially at the molecular level, in order to eventually leverage the diagnostic and therapeutic potential of the microbiome. Experimental work is needed to validate our findings, and future studies would benefit from whole (meta-)genome sequencing and fecal metabolite quantification (perhaps in parallel with serum), to better disentangle potential host and microbial contributions to inflammatory disease states^61^. More end-to-end collaboration between clinicians, experimentalists, and statisticians is needed to design studies which integrate molecular -omics approaches to understanding disease mechanisms^62–64^ with established diagnostic and treatment criteria, and biomarkers like fecal calprotectin and serum zonulin^65,66^.

## Supporting information

SFig. 1A, SFig. 1B, SFig. 2, Supplementary Methods, Supplementary Table

## Data Availability

All data produced are available online at https://github.com/sxmorgan/gespic-public

## Data Availability

Processed data tables mentioned in the supplemental file are hosted at https://github.com/sxmorgan/gespic-public. All software and R package versions are denoted in the *renv.lock* file also available in the repository.

## Funding Acknowledgements

This work was funded in part by the Deutsche Forschungsgemeinschaft (DFG, German Research Foundation) as part of a clinical research unit (CRU339): Food allergy and tolerance (FOOD@) - Project No 428046232) and CRC1449 to S.K.F., as well as CRC-TRR241 and CRC1449 to B.S. T.S. is supported by the Deutsche Forschungsgemeinschaft (DFG, German Research Foundation) under Germany’s Excellence Strategy – EXC 2155 “RESIST” – Project ID 390874280. Additional funding was received from the German Federal Ministry for Health and Research (BMBF) as part of the WHEAT-A-BAIC consortium, and from the Berlin Institute of Health (BIH). The OptiRef control cohort was partially supported from an unrestricted research grant from Novartis, and the AAU cohort by a grant from AbbVie. Dr. Judith Rademacher is a participant in the BIH-Charité Clinician Scientist Program funded by the Charité—Universitätsmedizin Berlin and the Berlin Institute of Health.

## Author Contributions

VRR and DP conceived the study and organized the experimental collection with help from JR, FP and UP. ME, VRR, BS, SKF, and DP developed the hypotheses for the analysis. VRR prepared the stool samples which TS sequenced and ME processed with input from UL. ME and VRR, JM and JK designed the metabolomics pilot study. VRR prepared the serum samples which JM analyzed. ME conceived and performed the statistical analyses with input from VRR and SKF. ME and VRR interpreted the results with input from SKF, DP, and LM. ME produced the figures with guidance from VRR. VRR and ME wrote the manuscript. All authors discussed and approved the final manuscript.

