## Supplementary material for "Spondyloarthritis, acute anterior uveitis, and Crohn’s disease have both shared and distinct gut microbiota": SFig. 1A, SFig. 1B, SFig. 2, Supplementary Methods, Supplementary Table

Supplementary Data File

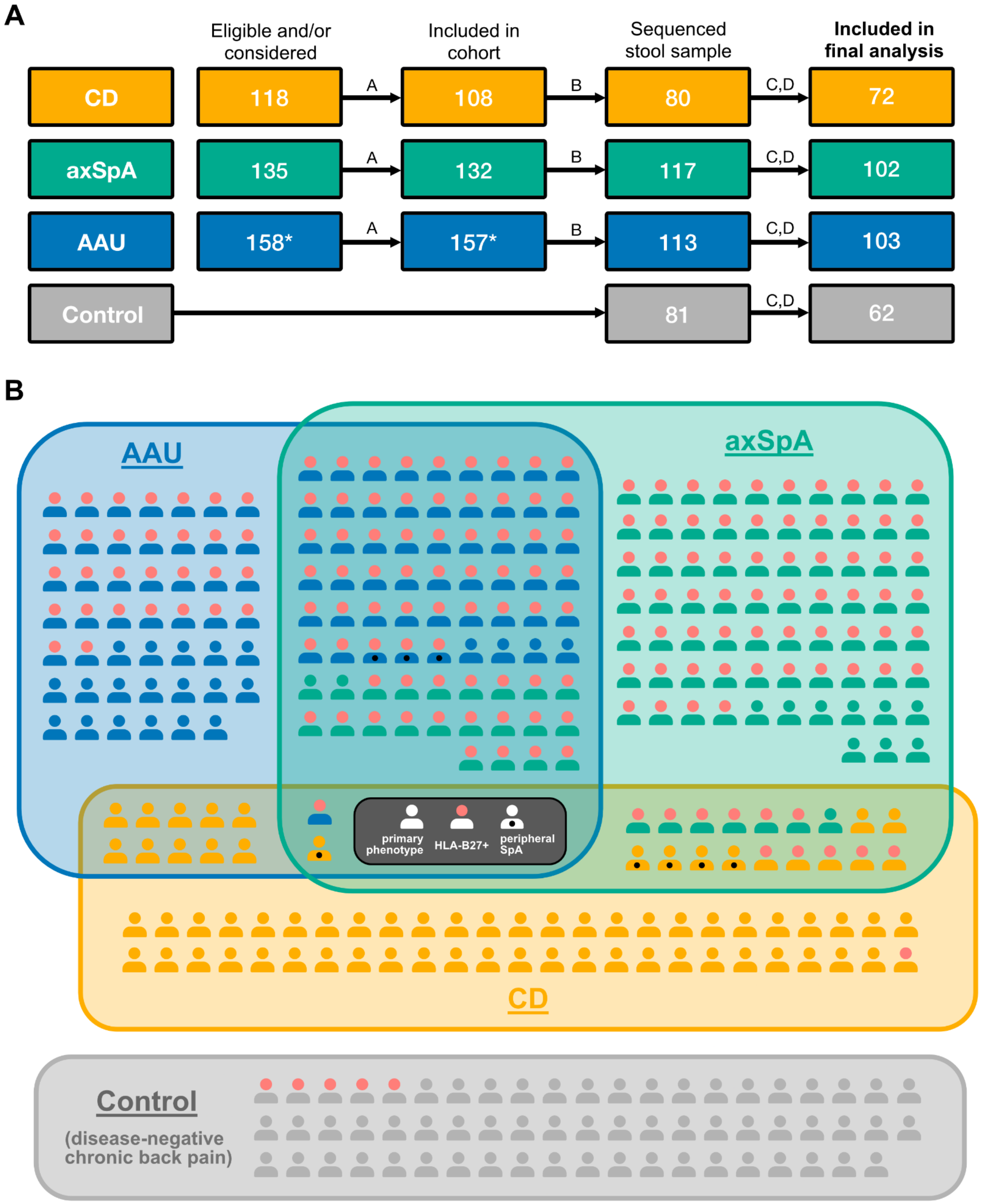


**Supplementary Figure 1: Overview of cohorts and individuals included in this study**
**A)** Flowchart containing numbers of participants at each stage of study with letters denoting reasons for exclusion, namely: *A*=excluded if patient inclusion criteria not met, *B*=excluded if no stool sample given at baseline visit, *C*=excluded if metadata incomplete or sequenced stool sample of poor quality, *D*=excluded if oral antibiotics taken ≤30 days prior to stool sample, or if previous diagnosis of SpA, AAU, CD, UC, or psoriasis present. An asterisk* denotes that recruitment was still ongoing at the time of analysis. Control samples were obtained from the Optiref study[1] and were resequenced and reprofiled with patient samples. **B)** Patients were recruited into cohorts reflecting their primary disease pathology, but varied in their expression of human leukocyte antigen (HLA) B27 and their presentation of secondary and tertiary pathologies. CD, Crohn's disease; axSpA, axial spondyloarthritis; AAU, acute anterior uveitis.

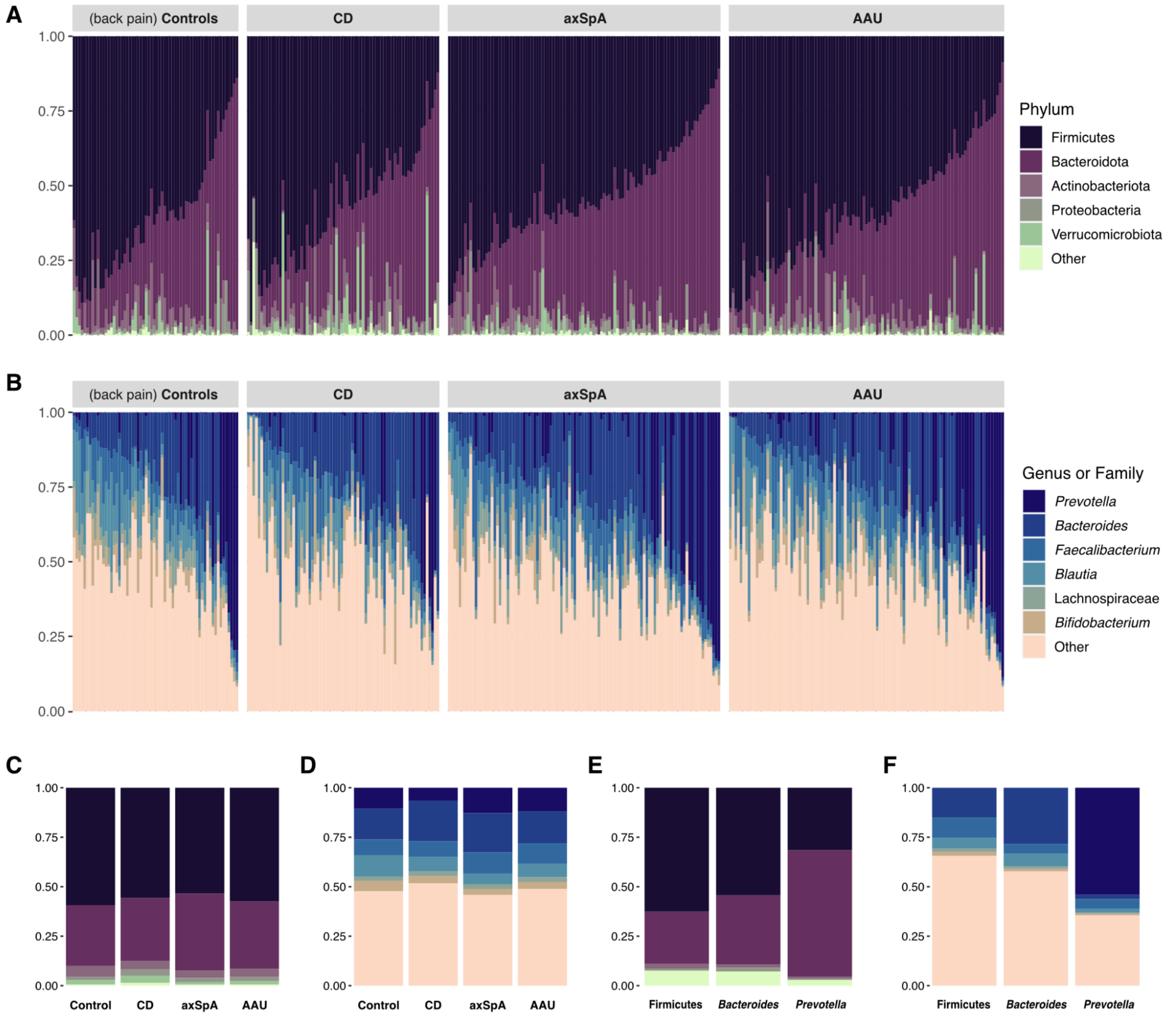


**Supplementary Figure 2: Microbiota composition of immune-mediated disease cohorts and back pain controls.** Relative abundances at different taxonomic resolutions of all n=339 individuals in our study, grouped by primary (recruitment) cohort. **A)** Phylum-level taxonomic profiles. Phyla with a median relative abundance of 0 across at least two cohorts were grouped into "Other". **B)** Taxonomic profiles of the top six most abundant taxa, corresponding to relative abundances binned at the genus level, except for Lachnospiraceae (representing an unknown genus inside the Lachnospiraceae family). All other taxa were pooled into "Other". Both legends are ranked according to median relative abundance across all cohorts from top to bottom and samples follow the same order in A and B. **C)** Median relative abundances of phyla by cohort. **D)** Median relative abundances of genera by cohort. **E)** Same as C but for enterotype groups determined from the bins in D (modification from Lahti and Shetty[2], see main text **Methods**), representing the sum of all ASVs bearing the indicated genus label. **F)** Same as D, but as in E shown for each enterotype.

All supplementary tables are hosted at www.github.com/sxmorgan/gespic-public, and printed here if space allows.

**Supplementary Table 1: Taxonomic annotations for all n=442 ASVs used in the differential abundance analysis**. Species annotations were inferred using the *dada2::addSpecies()* function with *allowMultiple=F*, in order to exclude identification of ASVs where a species-level consensus was not reached[3]. For the binned differential abundance analysis, the column "bin" was used to sum ASV rows.

| Covariate | Variable type | Description |
|---:|---|---|
| GESPIC | Binary | Any disease diagnosis |
| Age | Continuous | Age in years |
| Sex | Binary | Sex assigned at birth |
| BMI | Continuous | Body mass index |
| HLAB27 | Binary | HLA-B27 expression |
| CRP | Continuous | C-Reactive Protein in mg/L |
| ESR | Continuous | Erythrocyte sedimentation rate in mm/h |
| BASDAI | Ordinal | Bath Ankylosing Spondylitis Disease Activity Index |
| SpA | Binary | Previous or current spondyloarthritis diagnosis |
| CD | Binary | Previous or current Crohn's diagnosis |
| AAU | Binary | Previous or current acute anterior uveitis diagnosis |
| PsA | Binary | Previous or current psoriatic arthritis diagnosis |
| Antibiotics_recent | Binary | Antibiotic treatment 2-3 months before sampling |
| Corticoid_current | Binary | Current glucocorticoid treatment |
| csDMARD_ever | Binary | Current conventional synthetic DMARD treatment |
| csDMARD_current | Binary | Previous conventional synthetic DMARD treatment |
| NSAID_current | Binary | Current NSAID treatment |
| Smoker | Binary | Current smoker |
| Cohort_CD | Binary | Crohn's disease cohort (primary CD diagnosis) |
| Cohort_SpA | Binary | Axial spondyloarthritis cohort (primary SpA diagnosis) |
| Cohort_AAU | Binary | Acute anterior uveitis cohort (primary AAU diagnosis) |
| LibSize | Continuous | Raw number of sequencing reads from stool sample |

**Supplementary Table 2: Complete list of tested covariates available for all patients and controls**
GESPIC: German Spondyloarthritis Inception Cohort; HLA-B27: human leukocyte antigen B27; DMARD: disease-modifying anti-rheumatic drug; NSAID: non-steroidal anti-inflammatory drug

**Supplementary Table 3: Master table of all n=339 anonymized individuals in our study.** ASV and higher taxa (binned at genus- or family-level) relative abundance profiles, alpha diversities, clinical metadata (NA if missing), and serum metabolite concentrations (NA if below limit of detection) are included in wide format.

### Supplementary Methods

**Targeted Metabolomic Analysis**

We opted to quantify serum samples because fecal samples are difficult to sample consistently and reflect what is being expelled from the gut, not what the body has access to. In addition, there are standard reference materials (SRMs) that can be used for consistent quality management across batches and studies of serum. A subset of 48 serum samples were selected for tryptophan and SCFA metabolite analysis based on random subsampling within cohorts (n=12 from each of the disease groups plus controls) and using a seed for reproducibility. Relevant constraints were used for each cohort to ensure proportions of key variables observed in the larger dataset were maintained (i.e. HLA-B27 expression, disease activity, and enterotype). One serum sample was mislabeled during sample processing and could not be used for further analysis.

Two quantitative UPLC-MS/MS analyses were carried out independently using dedicated methods that were validated following European Medicines Agency (EMA) guidelines[4]. For each, metabolite extraction was performed followed by injection on a UPLC system 1290 Infinity II (Agilent, Santa Clara, CA, USA) coupled to a TSQ Quantiva (ThermoFisher Scientific, Waltham, MA, USA). For these analyses, commercial standards were spiked and then serially diluted into charcoal single stripped serum (Innovative Research, Novi, MI, USA) to create a calibration curve for quantification, along with solvent blanks and processed blanks. For quality management, Quality Control (QC) samples consisting of pooled samples and charcoal stripped serum samples spiked with a mix of commercial standards at known concentrations (QC standards) were also regularly injected for each batch.

The analysis of SCFA and ketone bodies (n = 11 metabolites in total) followed a previously published method[5]. In a glass vial, 40 µL of sample was incubated for 45 min at 40°C with 20 µL of 200 mM 3-Nitrophenylhydrazine hydrochloride, 20 µL of 120 mM N-(3-Dimethylaminopropyl)-N′-ethylcarbodiimide hydrochloride with 6 % pyridine, and 10 µL of Acetonitrile:Water (50:50) (v/v) containing stable isotope labeled internal standards (ISTDs) at 50 µM. This was followed by the addition of 410 µL of ACN:$H_2O$ (10:90) (v/v) and centrifugation at 5,500 x *g* for 20 min at 20° C. The supernatants were then transferred to HPLC vials with inserts for UPLC-MS/MS analysis.

Liquid chromatography used a RP C18 column ACQUITY BEH 1.7 µm 2.1x100mm (Waters Corp, Milford, MA, USA) at 40 ºC for separation, with $H_2O$ + 0.1 % (v/v) formic acid (FA) and ACN + 0.1 % (v/v) FA as Phase A and Phase B, respectively. A 0.35 mL $min^{-1}$ flow was maintained and the following gradient was used: 5 % B for 5 min, 5-55 % B in 12 min, 100 % B for 1 min, and 2 min 5 % B. Injection volume was 5 µL.

The analysis of tryptophan metabolites was carried out according to an in-house developed protocol (Brüning *et al*, in final preparation). Briefly, in an Eppendorf tube, 50µL of matrix was extracted with 100 µL Methanol:Water (90:10) (v/v) with 0.2% FA and 0.02% ascorbic acid containing ISTDs at 0.15 µg/mL. After one-hour incubation at -20°C to allow protein precipitation, the samples were then centrifuged at 12000 rpm for 10 minutes at 10°C. Supernatant was then transferred to HPLC vial inserts for UPLC-MS/MS analysis.

A C18 reverse phase column (Waters XSelect Premier HSS T3 Column, 100Å, 2.5 µm, 2.1 X 150 mm) was used at 15°C with the $H_2O$ + 0.2 % (v/v) FA (Phase A) and Methanol + 0.2 % (v/v) FA (Phase B). The following elution gradient was carried out at 0.4 mL $min^{-1}$: 3% B for 0.45 min, 30% B at 1.2 min, 60% B at 2.7-3.75 min, 95% B at 4.5-6.6 min, 3% B at 6.75-10 min. 5µl were injected per sample.

For both methods, data were then processed with Skyline where metabolite peaks were manually identified and integrated. Area under the curve (AUC) results were exported and processed using the dedicated in-house R package *MetaProc* (paper in preparation). Each analyte was normalized to its respective internal standard, followed by drift correction using the *notame* package[6], background correction for charcoal stripped serum samples and concentration calculation based on the calibration curve. Precision (RSD <15%) and accuracy (-/+ 15% theoretical concentration) were checked using the QC pools and QC standards. Then, for each measured sample, different statuses were automatically assigned according to calculated Limit of Detection (LOD), Lower and Upper Limits of Quantification (LLOQ, ULOQ, respectively). When the calibration curve fit ($R^2$>98%), precision and accuracy did not match previously defined thresholds and/or when the signal was lower than LOD in all samples, the analyte was not further considered.
For subsequent statistical analysis, concentration results were used only when the analyte could be quantified between LLOQ and ULOQ. When the analyte signal was detected between LOD and LLOQ for most samples, only normalized data was retained for relative comparisons. To reveal correlations with host phenotypes and microbial taxa of interest, Wilcoxon or Spearman tests were performed with valid concentration measurements while blocking for relevant covariates using the *coin* R package. Significant associations ($q$<0.05) between metabolites and the shared phenotype were independent of HLA-B27 status and each of the dichotomous phenotype labels (CD, AAU, SpA).
